# COVID-19 vaccination was a rare potential etiology for cause of death after medicolegal autopsy. A Finnish nationwide study

**DOI:** 10.1101/2024.02.13.24302742

**Authors:** Lasse Pakanen, Tuomo Nieminen, Paula Kuvaja, Hanna Nohynek, Sirkka Goebeler, Miia Artama, Petteri Hovi

## Abstract

COVID-19 vaccinations began globally at the end of 2020. By the end of 2021, 9.8 million doses were given in Finland. Regarding safety, most vaccine-related adverse reactions have been mild, but serious and lethal ones have also occurred. Autopsies in post vaccination deaths may give insight to the extent of fatal health conditions with potential COVID-19 vaccine etiology and provide new hypotheses of possible causalities between vaccination and severe health conditions. We searched the complete documentation on all medicolegal autopsies in Finland between December 2020 and December 2021 to assess how often the basis for autopsy was a suspected fatal adverse reaction to COVID-19 vaccination, and whether vaccination remained a potential etiology for any health condition determined as a cause of death after the autopsy. We linked register-based data on individual COVID-19 vaccination course and pre-existing health conditions.

We found 428 autopsy cases with a mention of COVID-19 vaccination, and prior to autopsy, vaccination was suspected to play a part in 76 deaths. Post autopsy, a forensic pathologist considered vaccination as a potential etiology in five underlying and seven contributory causes of death. These included seven thromboembolisms, two diabetic ketoacidoses, one myocarditis, one acute pancreatitis, and one eosinophilic granulomatosis with polyangiitis. In relation to the number of vaccinations within Finland, a suspicion of vaccine-related serious adverse reaction was rarely an indication for medicolegal autopsy. Even less frequently was vaccination considered to play a part in the process leading to death, although considerable doubt remains in the accuracy of individual considerations, and autopsy cannot definitively confirm causality between vaccination and death. Regarding vaccination safety, continuing evaluation of suspected vaccine-related deaths is essential, and an autopsy should be part of the investigation when such a suspicion arises.

## INTRODUCTION

During the COVID-19 pandemic, fast development of vaccines created several products that efficiently prevent severe illness and death caused by the SARS-CoV-2 virus (1). The vaccines were proven to be safe enough to meet the agreed emergency use licensing criteria, and large-scale as well as long-term safety follow-up was to be done only post licensure (2). Post-marketing safety surveillance has revealed rare severe adverse events following immunization (AEFI, terminology used in this paper according to the European Medicines Agency guideline (3), is presented in Table 1), with some considered to be truly caused by the vaccines. The first serious adverse reactions were thromboembolic complications including cerebral venous sinus thrombosis (CVST) and other unusual thrombotic events which were related to adenoviral vector vaccine ChAdOx1 (Vaxzevria, AstraZeneca) as reported by European Medicines Agency in April, 2021 (4). Subsequently, an association emerged between these thromboses and thrombocytopenia, as well as the occurrence of anti-platelet factor 4 antibodies in the patients, and this syndrome was named vaccine-induced immune thrombotic thrombocytopenia (VITT) (5), or thrombosis with thrombocytopenia syndrome (TTS) (6). Notably, VITT is also considered an adverse reaction to another adenoviral vector vaccine, Ad26.COV2.S (COVID-19 Vaccine Janssen, Janssen Group) (5, 7). Additionally, cases of myocarditis and pericarditis have occurred shortly after vaccination, and these conditions have been shown to be associated with mRNA vaccines BNT162b2 (Comirnaty, BioNTech-Pfizer), and mRNA-1273 (Spikevax, Moderna) in several large epidemiological studies, especially in young men (8–10). However, though myocarditis is a severe condition, reports in young men show relatively mild outcomes and no mortality associated with myocarditis shortly after vaccination (11–12). Furthermore, based on a population-based follow-up study of vaccine-related myocarditis, deaths are very rare (13).

**Table 1.** Definitions of vaccine-related incidents used in this paper.

| **Term** | **Definition** |
| --- | --- |
| Adverse event following immunization (AEFI) | Any health condition or problem that occurs following immunization and which does not necessarily have a causal relationship with vaccination (3) |
| (Serious) adverse reaction | Noxious and unintended health condition in response to vaccination with a suspected causal relationship. Serious adverse reactions include health conditions which are life-threatening or result in death (3). |
| Unrelated health problems following vaccination | Health condition considered not to be caused by vaccine |
| Cause of death | Cause of death as stated in the death certificate determined by a forensic pathologist or another physician |

A vast amount of unconfirmed and misinterpreted data without epidemiological support regarding vaccine-related morbidity and mortality has been of great concern from the beginning of COVID-19 vaccinations (14–15). For an individual who dies within days or weeks following vaccination, the causes of death should be thoroughly examined on one hand because of individual legal protection, and, on the other hand, to reveal potential severe health conditions following immunization. The key investigation in this regard is an autopsy, either medicolegal or clinical. A growing number of autopsy case reports of deaths following COVID-19 vaccination have been published (e.g., systematic review by Sessa et al. (16)). However, few larger autopsy-based data have yet been published. The Finnish law requires that a medicolegal cause-of-death investigation must be done when death is considered unexpected, or if it has been caused or suspected to have been caused by non-natural causes such as medical treatment (Act on the Inquest Into the Cause of Death 459/1973). Thus, a medicolegal autopsy is generally instructed by the police when a vaccination is suspected to play a role in the process leading to death.

We set out to evaluate how often the basis for medicolegal autopsy was a suspected fatal adverse reaction to COVID-19 vaccination in Finland, and we describe cases in which COVID-19 vaccination remained as a potential etiology for a health condition considered as any cause of death after the autopsy. We also describe those cases in which the forensic pathologist considered COVID-19 vaccination not to have any part in the death process. We describe the medicolegal cause-of-death investigation procedure and discuss the remaining uncertainties in both the accuracy of case-selection for assessment, and in determining the potential vaccine etiology.

## MATERIAL AND METHODS

### COVID-19 vaccinations in Finland

Since the beginning of the SARS-CoV-2-virus pandemic in 2020, several COVID-19 vaccines have been introduced to the market. In Finland, the first vaccines were given in December 2020. By the end of 2021, 9.8 million doses had been administered in the country (17), and nearly 90% of the population aged 12 years or older had received at least one dose (18).

Initially prioritized groups for COVID-19 vaccinations were the elderly, adults with a risk for severe COVID-19 disease, and health care workers. Vaccination was gradually extended during 2021 to cover the whole adult population with priority by age group from older to younger, and finally to children over five years old. By the end of 2021, five vaccine products had gained conditional marketing authorization: BNT162b2 on Dec 21, 2020; mRNA-1273 on Jan 6, 2021; ChAdOx1 on Jan 29, 2021; Ad26.COV2.S on Mar 11, 2021; and NVX-CoV2373 (Nuvaxovid, Novavax) on Dec 20, 2021 (19). Due to safety concerns related to coagulation disorders, the use of ChAdOx1 ceased on Mar 19, 2021 for those younger than 65 years, and altogether on Nov 30, 2021 (20). Because of the suspected relationship of mRNA-1273 and myocarditis and pericarditis, the use of this vaccine ceased for men under 30 years of age on Oct 7, 2021 (21). All vaccinations given in Finland are recorded in electronic health record systems and collected to the National Vaccination Register (22).

### Medicolegal cause-of-death investigation process in suspected vaccine-related deaths in Finland

If a suspicion of a relationship between vaccination and death arises, the police instruct a medicolegal autopsy. Suspicions can be based on information from next-of-kin, from treating clinicians, from police investigation and/or in consultation with a forensic pathologist, and they can be fully subjective, based on clinical history, or other circumstances prior to death, e.g. sudden unexpected death close to vaccination with or without a mention of adverse event. The process needs to be actively initiated and the suspicion can be brought forward by any of the mentioned parties. Usually this happens when death occurs close to vaccination, unexpectedly, or when no other obvious cause of death can be determined based on available clinical data. Threshold for initiating the medicolegal process is kept low especially when a relative expresses the suspicion (23). Finland has one of the highest autopsy rates among high income countries (24), with medicolegal autopsies carried out in 13.7% during the study period (Table 2; 25), and the overall autopsy rate being 17.6% of all deaths in 2021 (26).

**Table 2.** Medicolegal autopsies carried out in relation to total death during the study period, Dec 26, 2020 – Dec 31, 2021.

|  | **Medicolegal autopsies** | **Total deaths (25)** | **Autopsy-percentage** |
| --- | --- | --- | --- |
| End of follow-up | Dec 31, 2021 | Dec 31, 2021 |  |
| Number of cases | 8,048 | 58,714 | 13.7 |
| Sex |  |  |  |
| Men | 5,597 | 29,697 | 18.9 |
| Women | 2,451 | 29,017 | 8.5 |
| Age, years |  |  |  |
| < 18 | 80 | 214 | 37.4 |
| 18–34 | 531 | 646 | 82.2 |
| 35–64 | 2,751 | 6,862 | 40.1 |
| ≥ 65 | 4,686 | 50,992 | 9.2 |

The Finnish Institute for Health and Welfare conducts all medicolegal autopsies in its Forensic Medicine Unit, geographically distributed in five locations covering the whole country. The institute keeps a medicolegal database for all data related to cause-of-death investigation including autopsy reports, death certificates, police reports, and other related documents.

### Individual assessment of causes of death in medicolegal autopsy

Along with a complete macroscopic examination, each autopsy case includes a routine analysis of histological samples. Most cases also include a toxicological analysis to rule out intoxication, or to assess the use of alcohol, narcotics, and medications. Based on individual consideration, the forensic pathologist may request further auxiliary investigations, such as neuropathology, microbiological analyses in suspected infections, or serum tryptase level in suspected anaphylaxis (e.g. 27 for further reference).

After all relevant investigations, the forensic pathologist determines the most likely *underlying cause of death, intermediate causes of death, the immediate cause of death, and contributory causes of death* to be recorded in death certificate.

Causes of death, including statements regarding causality, are always approximations based on a case-by-case assessment and hence involve uncertainty. In a case of suspected vaccine etiology, currently no exact analysis or procedure exists regarding the COVID-19 vaccines in use, upon which a fatal health condition could definitively be determined as to be caused by a vaccine dose given. When assessing vaccination as a potential etiology for any cause of death, the forensic pathologist should consider several questions including, as suggested by the World Health Organization (28): 1) is there strong evidence for other causes; 2) is there a known causal association with the vaccine or vaccination; and 3) was the event within the time window of increased risk? Regarding the second and third questions, one must refer to the current, constantly developing scientific knowledge when determining vaccine as an etiology for any given health condition.

Likewise, no exact analysis exists upon which vaccination could with certainty be excluded as an etiology for a given health condition. Notably, a forensic pathologist must largely rely on published data, mostly epidemiologic, when assessing the causal connection between vaccination and death.

### Selection of medicolegal autopsy cases for further examination

Data in the medicolegal database are structured and include sex, age, causes of death, the date and location of death, and the personal identification code, which is given to all permanent residents of Finland upon birth or immigration. We examined all medicolegal autopsy cases with both the date of death and the date of autopsy between Dec 26, 2020 (the start of vaccinations in Finland) and Dec 31, 2021 (8,048 autopsy cases). We first excluded cases in which homicide, accident, suicide, or occupational disease was the only grounds for medicolegal autopsy. Then, we also excluded from further evaluation all cases with no mention of having received COVID-19 vaccination in any document recorded in the medicolegal database (police report, autopsy referral, any attached patient records, other related documents, or notably also the autopsy report and death certificate). From among the remaining cases in which vaccination was mentioned, one of the researchers, a forensic pathologist (L.P.), determined, whether an actual suspicion of a relationship between vaccination and death had arisen prior to autopsy based on the recorded data.

### Linkage to vaccinations data and risk factors from the national population-wide registers

We linked data between registers and autopsy cases using the personal identification code. We report the sex, age, and vaccination characteristics of the autopsied individuals, along with data from all vaccinated individuals in Finland for comparison. For this purpose, we identified all individuals, whose last known permanent address was in Finland according to the Population Information System, and who received COVID-19 vaccinations according to the Finnish National Vaccination Register between Dec 26, 2020 and Dec 31, 2021 (22). The vaccination register includes the date and vaccine brand of each given COVID-19 vaccination.

For descriptive purposes, we also linked our cases, along with those COVID-19 vaccinated based on the vaccination register, with data from the care register for health care and the medical reimbursements register on pre-existing risk factors predisposing to severe COVID-19 disease as described in the study of Salo et al. (29). These risk factors, together with age group, were important determinants of the priority and the number of recommended vaccinations for individuals in Finland, and they are also risk factors of death from any cause. These risk factors were: cardiovascular diseases, cognitive disorders, other neurological disorders, diabetes, chronic kidney diseases, chronic liver diseases, pulmonary diseases, immunosuppressive diseases, Addison’s disease, schizophrenia, and Down syndrome. Furthermore, data on 24-hour service housing and institutional care were gathered from the same registers. We gathered this risk data starting from Jan 1, 2015 until the first vaccine dose, or until death if no record of vaccination was found in the register. Medical reimbursement data was available starting from Jan 1, 2018.

### Ethics

This descriptive study was conducted as part of the Finnish Institute for Health and Welfare’s responsibility to monitor the safety of vaccines used in the national vaccination program and to investigate potential adverse events suspected to be linked with vaccination as dictated by the Finnish law (Act on the National Institute for Health and Welfare 668/2008; Government Decree on the National Institute for Health and Welfare 675/2008; and the Communicable Diseases Act 1227/2016). Based on this, Finnish Institute for Health and Welfare has the statutory right, notwithstanding confidentiality provisions, to access necessary information in cause-of-death investigation documents and to link this information with other relevant register data. Accordingly, this study was approved by the Finnish Institute for Health and Welfare’s Director of Health Security department, Director of Government Services department, as well as the Deputy Director General of Research, Development and Innovation. The data on the medicolegal autopsies was originally collected according to the Act on Determining the Cause of Death (459/1973).

## RESULTS

A total of 428 autopsy cases with a mention of having received COVID-19 vaccination were found, and their records were further investigated for the current report by a forensic pathologist (L.P.). A suspicion of vaccine etiology arose in 76 cases prior to autopsy (Figure). After the autopsy and auxiliary investigations, 12 cases emerged, in which the forensic pathologist who wrote the death certificate considered the COVID-19 vaccination as a potential etiological factor for any cause of death: in five cases for the underlying cause of death, and in seven cases for a contributory cause of death (Table 3). Three of these cases had no pre-autopsy suspicion of vaccine etiology, and, thus, were not part of the 76 cases (Figure).

**Figure.**
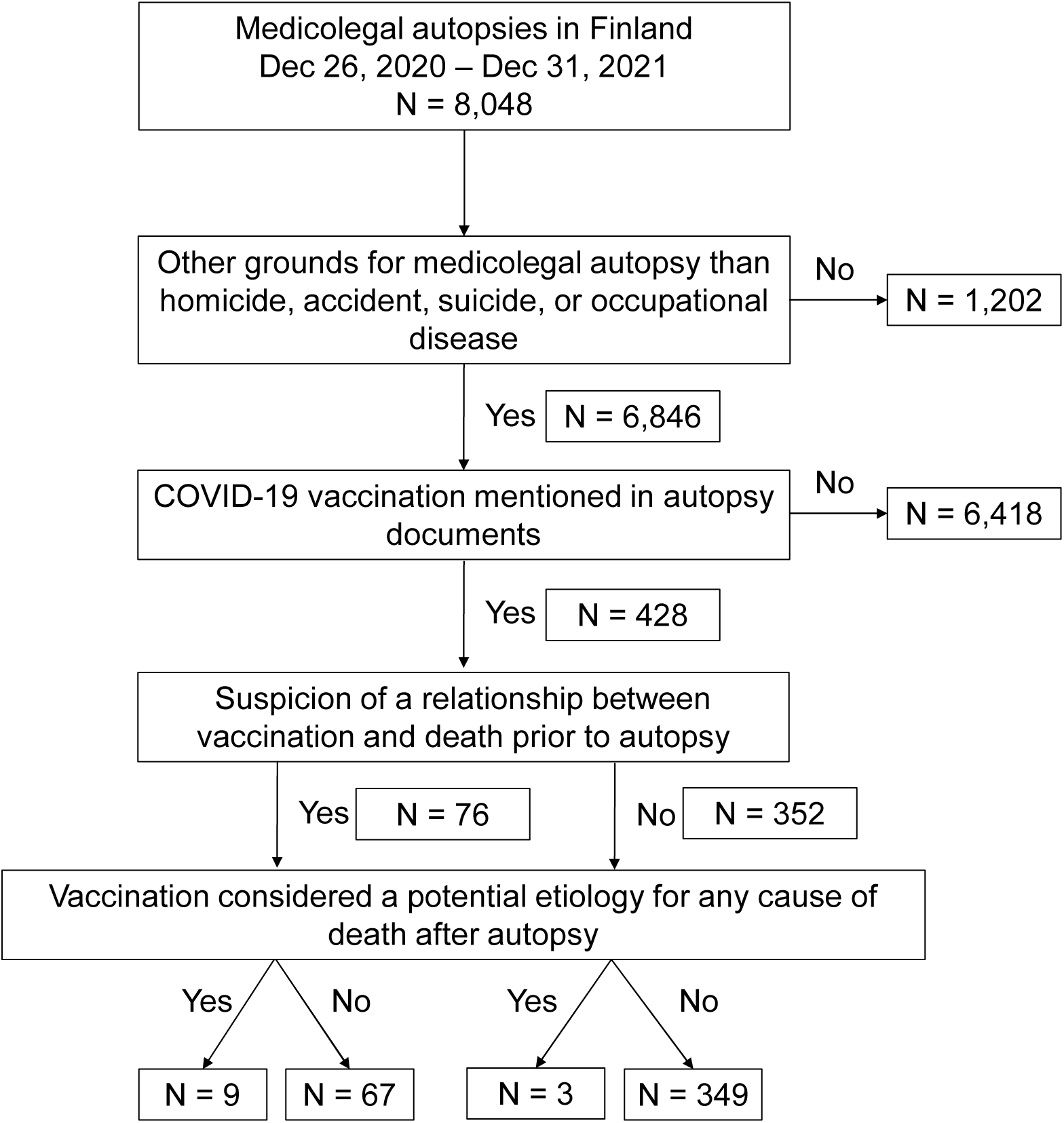
Flow chart presenting the selection of autopsy cases. Vaccination data was obtained from Finnish National Vaccination Register.

**Table 3.**
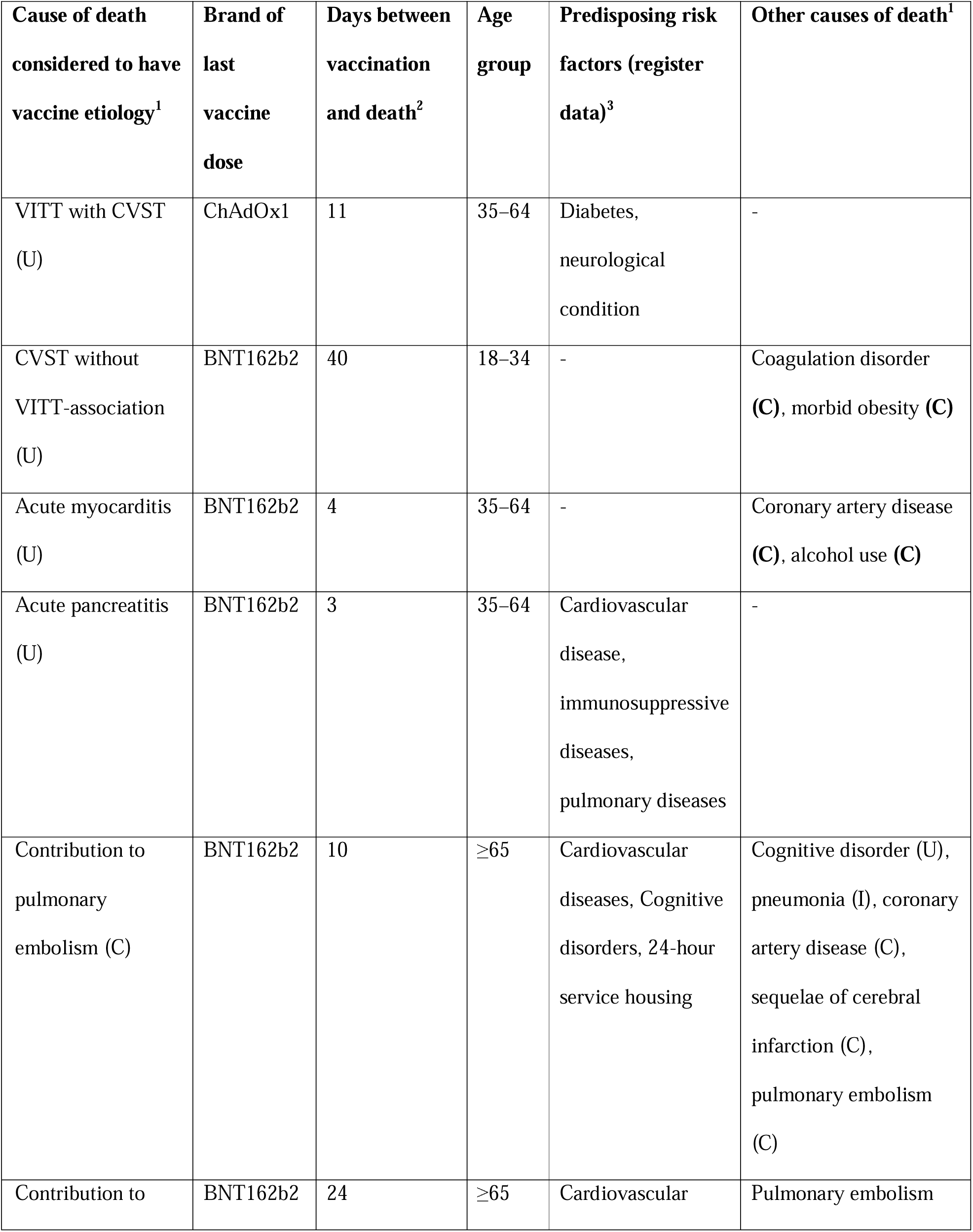
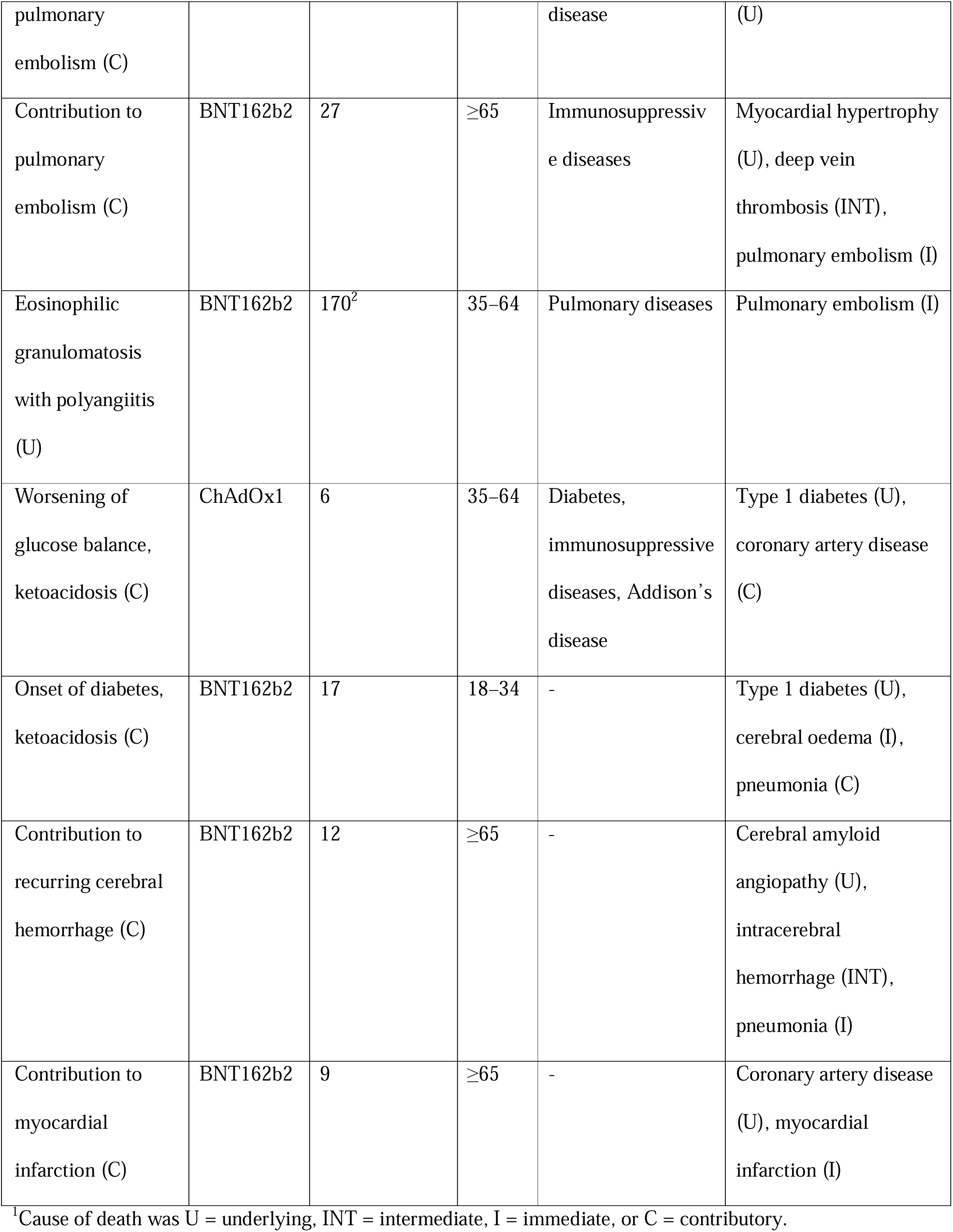
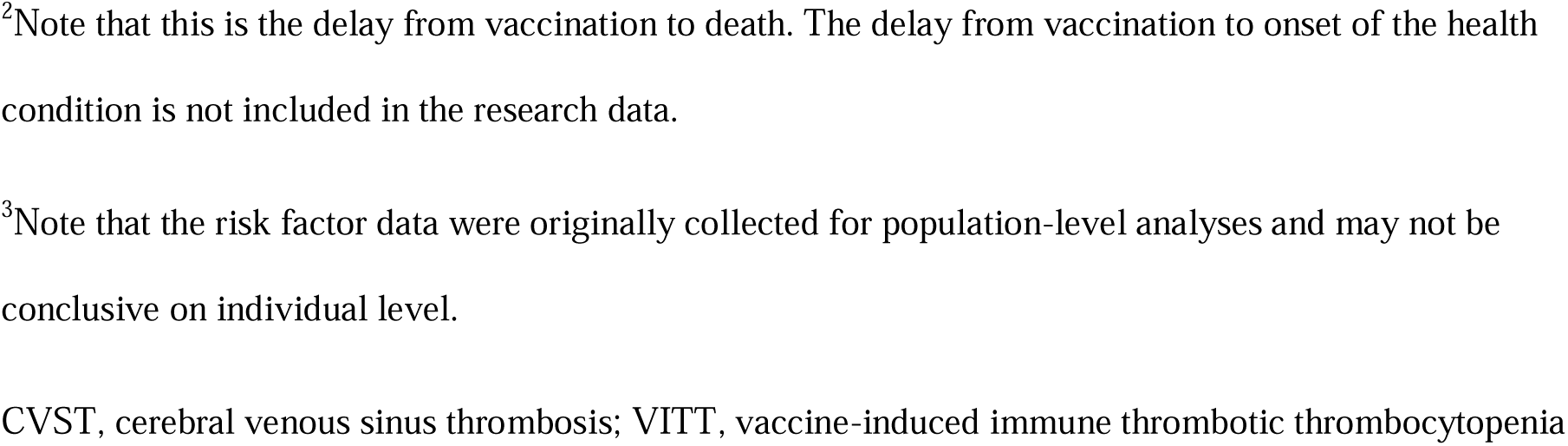
Details of all cases in Finland in which a forensic pathologist considered a cause of death to have COVID-19 vaccine etiology.

Details of the vaccinations preceding death existed in the Vaccination register for 417 cases, including all the 12 cases with potential vaccine etiology. In further 4 cases, the medicolegal data included a mention of vaccination with details, and in 7 cases, only a mention of vaccination without any details.

Most causes of death with potential vaccine etiology (n = 12, see Table 3) were thromboembolic events (7/12) including one VITT with CVST, and one CVST without VITT-association, three cases of pulmonary embolisms, one case of acute myocardial infarction, and one case of recurring cerebral hemorrhage. Furthermore, potential vaccine etiology arose in one case of acute myocarditis, in two cases of diabetic ketoacidosis, in one case of acute pancreatitis, and in one case of eosinophilic granulomatosis with polyangiitis (EGPA). Death occurred within 14 days after vaccination in 7/12 cases. The deaths without a considered vaccine etiology (67/76) were mostly caused by diseases, mainly of cardiovascular origin (Table 4).

**Table 4.** Causes of death in cases with suspected relationship between COVID-19 vaccination and death prior to autopsy but with no vaccine etiology considered after medicolegal autopsy.

| Cause of death | Number of cases (%) |
| --- | --- |
| Disease | 62 (92.5) |
| Cardiovascular diseases | 43 (64.2) |
| • Acute myocardial infarction | 9 (13.4) |
| • Arrhythmia | 1 (1.5) |
| • Other / unknown mechanism | 33 (49.3) |
| Cerebrovascular diseases | 2 (3.0) |
| Alcohol-related diseases | 1 (1.5) |
| Gastroenterological diseases | 3 (4.5) |
| Malignancies | 2 (3.0) |
| Pulmonary embolism | 5 (7.5) |
| Pulmonary diseases | 2 (3.0) |
| Endocrinopathies and nutritional diseases | 2 (3.0) |
| Neurodegenerative diseases and dementia | 2 (3.0) |
| Accident | 4 (6.0) |
| Undetermined | 1 (1.5) |
| Total | 67 (100.0) |

Prior to autopsy, the suspicion of a relationship between vaccination and death came from treating clinicians in 28 (36.8%) cases, a relative in 24 (31.6%) cases, forensic pathologist in 8 (10.5%) cases, and the police in 1 (1.3%) case (more than one source was possible in each individual case). Twenty-two (28.9%) cases had a mention of a health condition preceding death suspected to be a vaccine-related (mild) adverse reaction, but no actual suspicion of it causing death was stated. Death occurred at home in 48 (63.2%), in a health care unit in 16 (21.1%), in a social welfare unit in 5 (6.6%), and in other places in 7 (9.2%) cases. As regards the 12 cases with potential vaccine etiology, the location of death was home in five cases, health care unit in six cases, and other in one case.

The 12 deaths with potential vaccine etiology occurred in all adult age groups and equally in both sexes (Table 5). Half the cases were autopsied in Helsinki, which has proportionally the most autopsies, and all five autopsy locations had at least one case. Four deaths occurred after each first, second, and third dose, while the number of total administered COVID-19 vaccinations were 4.3, 4.1, and 1.3 million, respectively. Ten of the 12 deaths (83%) occurred after vaccination with BNT162b2, which was the vaccine brand with the largest number of vaccinations administered out of all vaccinations (7.9M / 9.8M; 81%). All 12 cases with potential vaccine etiology had one or more chronic health conditions according to the person level register linkage (Table 3): 7/12 (58%) cases had a record of at least one known pre-existing risk factor, which was a larger proportion than with those vaccinated (1.0M / 4.3M; 23%, Table 5). In 8/12 cases, including the remaining five cases without a register record of risk factors, forensic pathologist determined at least one corresponding chronic condition as one of the causes of death after the autopsy.

**Table 5.** Data on demographics, vaccinations, and pre-existing health conditions.

|  | **COVID-19 vaccine considered as etiology for any cause of death after medicolegal autopsy** | **Suspicion of a relationship between COVID-19 vaccine and death prior to medicolegal autopsy** | **Exposed population** |
| --- | --- | --- | --- |
| End of follow-up | Death | Death | Dec 31, 2021<sup>1</sup> |
| Number of cases | 12 | 76 | 4,345,346 |
| Sex |  |  |  |
| Men | 6 | 34 | 2,100,677 |
| Women | 6 | 42 | 2,244,669 |
| Age, years |  |  |  |
| < 18 | 0 | 0 | 323,599 |
| 18–34 | 2 | 4 | 930,650 |
| 35–64 | 5 | 23 | 1,888,942 |
| ≥ 65 | 5 | 49 | 1,202,155 |
| Most recent vaccine dose of the deceased vs. population vaccinated with cumulative number of doses |  |  |  |
| 1 | 4 | 45 | 4,345,346 |
| 2 | 4 | 19 | 4,135,987 |
| 3 | 4 | 9 | 1,265,677 |
| 4 | 0 | 0 | 3,042 |
| Vaccinations |  |  |  |
| ChAdOx1 | 2 | 19 | 553,426 |
| BNT162b2 | 10 | 46 | 7,909,387 |
| mRNA-1273 | 0 | 8 | 1,285,369 |
| Ad26.COV2.S | 0 | 1 | 206 |
| Other | 0 | 0 | 1,664 |
| Total |  |  | 9,750,052 |
| Number of pre-existing risk factors<sup>2</sup> |  |  |  |
| 0 | 5 | 19 | 3,328,945 |
| 1 | 3 | 15 | 804,011 |
| 2 | 2 | 17 | 176,518 |
| ≥3 | 2 | 25 | 35,872 |
| Nursing home residency | 0 | 1 | 28,763 |
| Need for 24-hour care | 1 | 10 | 65,532 |
| <sup>1</sup> Date for calculating age at first dose.<br><sup>2</sup> Pre-existing risk factors up until the first given vaccine dose, (each as 0 or 1): cardiovascular diseases, cognitive disorders, other neurological disorders, diabetes, chronic kidney diseases, chronic liver diseases, pulmonary diseases, immunosuppressive diseases, Addison's disease, schizophrenia, and Down syndrome |  |  |  |

## DISCUSSION

In this nationwide analysis, we searched 8,048 medicolegal autopsy reports comprising all medicolegal autopsies in Finland from Dec 26, 2020 to Dec 31, 2021. For comparison, 58,714 deaths from all causes occurred during this time (Table 2; 25), among which were 952 deaths caused by COVID-19 infection in 2021 (30). Moreover, 9.8 million COVID-19 vaccinations were given in Finland, and Finnish Institute for Health and Welfare has estimated that the vaccinations prevented more than 7,300 deaths from COVID-19 disease and more than 1,000 deaths with COVID-19 disease as a contributory factor in the country by the end of March, 2022 (31). According to our data, 76 cases included a pre-autopsy suspicion of a relationship between COVID-19 vaccination and death, and, after the autopsy, forensic pathologist considered COVID-19 vaccination as a potential etiology for five underlying, and seven contributory causes of death, mainly from thromboembolic causes (7/12). In comparison to the total vaccinated population, the 12 deaths with potential vaccine etiology occurred in both sexes and all adult age groups but included more of those with a third dose and those with known pre-existing risk factors. Due to the remaining uncertainties in catching the right cases, and in considerations about the etiology for the causes of death, the interpretations from the data need to be extremely cautious.

To the best of our knowledge, this is the largest published systematic case report series on autopsies conducted in deaths suspected to have COVID-19 vaccine etiology. Most autopsy studies have been single case reports as reviewed by Sessa et al. (16), and a growing number of similar anecdotal reports have been published since. In a Japanese study including 54 cases with death within seven days post vaccination that were autopsied, five cases were concluded to have possible causal relationship between vaccination and death (32). A study from Singapore included 29 autopsy cases from deaths within 72 hours after receiving mRNA-type vaccination, but they did not find any potential causative relationship between vaccination and death in these cases (33). Schneider et al. (34) reported 18 autopsy cases in which death occurred 1–14 days after vaccination in Germany. Of these, five deaths had a potential vaccine etiology including four cases of VITT and one myocarditis (34). Also in Germany, Schwab et al. (35) described 25 autopsy cases with death within 20 days post vaccination, of which five cases died from myocarditis with a potential mRNA-vaccine etiology. Chaves et al. (36) could not find any deaths potentially related to COVID-19 vaccination among the 121 autopsies of deceased that had received vaccination in Colombia. In the light of these case series studies, research is needed to reveal to what extent COVID-19 vaccine-related deaths do occur. However, no conclusions can be made on the frequency of potential vaccine-related deaths based on autopsy-studies alone, as the selection criteria for autopsy differ between studies and, more importantly, between countries.

In seven of the twelve deaths in our data, forensic pathologist writing the death certificate considered COVID-19 vaccination a potential etiological factor for a diagnosed thromboembolism, which were also the most frequent causes of death considered vaccine-related in previous reports (16). Among these seven cases, only one case of VITT with CVST was identified. Another CVST without thrombocytopenia was also detected. Due to the relatively early cessation of vaccination with ChAdOx1, exposure to this vaccine brand was a minority in Finland (Table 5), which may have limited the occurrence of these potentially lethal adverse reactions. The estimated cumulative 28-day-incidence of VITT has been between 1/26,500 and 1/1,000,000 first doses of ChAdOx1 with lower incidences for subsequent doses (5, 37–38). According to See et al. (39), the incidence of VITT after vaccination with Ad26.COV2.S was 1/583,000 doses. The estimated incidence for VITT with CVST after receiving the first dose of ChAdOx1 was 12.3 / 1,000,000 when the condition was first recognized as an adverse reaction (40). The rate is clearly higher than the background cumulative 28-day incidence of 0.18 / 1,000,000 (41). Mortality caused by VITT in the COVID-19 vaccinated is unknown, but it is thought to be extremely low (37). However, among patients with diagnosed VITT, the mortality rates have varied 23–73% (5), though with early recognition and adequate care, the prognosis has improved (42). Post vaccination CVST can occur with or without the context of VITT, but with significantly better outcomes, when no thrombocytopenia-association is present (40, 43).

Of the seven cases with thromboembolism, three deaths were from pulmonary embolisms, one death from acute myocardial infarction, and one death from recurring cerebral hemorrhage. Deep vein thrombosis and pulmonary embolism are common entities, and the latter is frequently the cause of sudden death in the medicolegal context. Case reports of post vaccination pulmonary embolisms have been published (44), but further population-based studies on ChAdOx (45), and BNT162b2 (46) found no evidence of any increased incidence of pulmonary embolisms after vaccination. However, in a self-controlled case series study in the French population, a slight increase in risk for pulmonary embolism was associated with the first dose of ChAdOx1 (relative incidence 1.41, confidence interval 1.13–1.75) but not with BNT162b2 or mRNA-1273 (47). Myocardial infarctions and other acute cardiovascular events are common manifestations in the general population, and they occur after vaccination as well. It has been suggested that COVID-19 vaccination could contribute to cardiovascular events through enhancing prothrombotic state or, unspecifically, as a stress factor (48). As with pulmonary embolism, Botton et al. (47) found an increase in the risk for myocardial infarction after the first dose of ChAdOx1 (relative incidence 1.29, confidence interval 1.11–1.51), and after Ad26.COV2.S (relative incidence 1.75, confidence interval 1.16–2.62), but not with the mRNA-type vaccines. Again, no increased risk for myocardial infarction arose in connection with BNT162b2 in other large study settings as reviewed by Yong et al. (49). Hence, although COVID-19 vaccination was, according to the medicolegal assessment, considered a potential etiological factor for pulmonary embolism in three of our cases and myocardial infarction in one case, all vaccinated with BNT162b2, we found no epidemiological evidence to support these decisions.

One case of post vaccination fatal acute myocarditis with potential vaccine etiology was present in our study population in the age group 35 to 64 years. A link between myocarditis and mRNA-type COVID-19 vaccines has been established in large cohort studies (10–11). The highest risk for developing myocarditis has occurred in young men after receiving the second dose (50) Case reports of vaccination-associated myocarditis deaths have been published (35, 51), but, in large study settings, post vaccination myocarditis has rarely been associated with death (8, 10, 13, 50, 52). Notably, myocardial injury associated with a clinical diagnosis of myocarditis after COVID-19 vaccination has been reported mild, as reviewed by Garg et al. (53). In the present study, forensic pathologist considered COVID-19 vaccination as contributory cause of death in two cases with type 1 diabetes mellitus and diabetic ketoacidosis. In one of them, the vaccine was a potential etiological factor for the onset of diabetes, and in the other, it potentially contributed to the worsening of glucose balance in a patient with previously diagnosed diabetes. Many case reports and case series have been published describing post vaccination hyperglycemia, hyperosmolar hyperglycemic syndrome, and diabetic ketoacidosis regardless of the COVID-19 vaccine type as reviewed by Samuel et al. (54). However, we could find only one self-controlled case series analysis regarding acute diabetic complications in patients with type 2 diabetes receiving BNT162b2 (55). According to the study of Wan et al. (55), no ketoacidoses emerged among the 141,224 cases given BNT162b2, and no increased risk for other diabetic complications was observed, either. We did not find any large-scale studies about the effects of COVID-19 vaccines on the onset or on the clinical picture of type 1 diabetes. These kinds of epidemiological studies are warranted, as in some case studies, COVID-19 vaccines were suggested to play a role in the onset of autoimmune diseases, including type 1 diabetes (as reviewed by Chen et al. (56)).

Herein, one case of fatal acute pancreatitis emerged with COVID-19 vaccination as its potential etiology. A few case reports about non-fatal post vaccination pancreatitis have been published, mainly concerning BNT162b2 (e.g., 57–58), but we did not find any epidemiological studies addressing the potential association of vaccination and pancreatitis. Lastly, we had one autopsy case who had EGPA, a small-vessel vasculitis, determined as the underlying cause of death, and COVID-19 vaccine considered as potential etiology for the disease. As with pancreatitis, only a few case reports of new onset or relapse of EGPA after COVID-19 vaccination have been published, all regarding mRNA vaccines (59–62). Our findings suggest that further research on the relationship between COVID-19 vaccination and both acute pancreatitis and EGPA is warranted.

As we have described in the methods, determining as well as excluding vaccination as an etiology for any cause of death involves a lot of uncertainty, and should be based on epidemiological evidence of association between the vaccination and the adverse event which contributed to the fatality (28). Also, it must be stressed that we only examined the outcomes of the cause-of-death investigation as done by each individual forensic pathologist, and, as such, we did not re-evaluate the possibility of vaccine-etiology in any of the cases. The medicolegal cause-of-death investigation process comprises of gathering information regarding patient history (known diseases, possible symptoms, and findings prior to death) and other circumstances preceding death, as well as examining data received from the autopsy and auxiliary investigations. Based on these, the forensic pathologist generates a most likely scenario including the causes of death and the potential etiologies involved. This scenario is then presented in the autopsy report along with discussion of the uncertainties in each case. It is possible that true vaccine-related deaths were missed, and some of these may even be among the 67 cases in which the forensic pathologist did not consider vaccination as potential etiology. Notably, among these were nine acute myocardial infarctions, five pulmonary embolisms, and one case in which the causes of death could not be determined. However, the lack of epidemiological support (47, 49), and the fact that more plausible etiologies were present (e.g. malignancy in case of pulmonary embolism) argue against a significant proportion of true vaccine-related deaths in our study.

Death occurred within 30 days of last vaccination in ten of the twelve cases with vaccination considered as an etiology for a cause of death. We did not examine the interval between vaccination and death in all included autopsies, as some authors have done (32–33, 35. All-cause mortality following COVID-19 vaccination has been studied on population level in Finland, and the hazard of death did not increase within 63 days of COVID-19 vaccination in Finland (63). Also, a meta-analysis including three self-controlled case series studies totaling 750,000 patients found no association between COVID-19 vaccination and all-cause mortality (64).

Because the initiation of the medicolegal process in a suspected vaccine-related death is dependent on a suspicion being brought up, not all cases end up in the hands of forensic pathologist, as seen in the context of iatrogenic deaths (65). This can more often be the case in deaths among elderly people with several chronic illnesses than in deaths deemed more unexpected, as is evident when comparing the medicolegal autopsy rates between 18-34-year-olds (82%) and 65-year-olds and older (9%).

Prior to autopsy, the suspicion of the role of COVID-19 vaccine in death was brought forward mostly by treating clinicians. Health care personnel are in a key position in spotting potential iatrogenic deaths, and they are required by law to report such cases to the police in Finland. The systems of investigating suspected iatrogenic deaths differ greatly between countries (66) depending partly on whether it is looked at from the viewpoint of clinical risk management, or whether actual malpractice claims have been raised (23).

In many cases, the suspicion of a relationship between COVID-19 vaccine and death was also expressed by the deceased’s next of kin, which is in line with today’s growing numbers of medical malpractice issues (67). COVID-19 vaccinations have been the topic of extensive public discussions globally, and views discouraging against vaccination on account of alleged hazards have occasionally gained ground (14). Because of this, it is essential to investigate all suspected COVID-19 vaccine-related deaths thoroughly and openly to address these concerns and, if so indicated, maintain the public’s confidence in vaccinations.

### Conclusions

In the context of total vaccinated population and of the number of given vaccine doses, only a few suspicions of vaccine-related deaths arose in the first year of COVID-19 vaccinations in Finland. Even fewer were the number of deaths, in which the forensic pathologist performing the cause-of-death investigation considered vaccine as a potential etiological factor for a cause of death. Although autopsy is a crucial tool in determining the causes of death through detection and documentation of tissue reactions, other settings, such as large epidemiological studies, should be performed to support individual case-assessment. Our study highlights the need for further research related to diabetes, acute pancreatitis and EGPA as potential serious adverse reactions of COVID-19 vaccines. Also, deaths associated with thromboembolic events and myocarditis after vaccination need closer examination. A continuing evaluation of suspected COVID-19 vaccine-related deaths is essential in monitoring vaccination safety.

## Data Availability

All data generated or analyzed during this study are included in this published article.

## Abbreviations

AEFI: adverse events following immunization
CVST: cerebral venous sinus thrombosis
TTS: thrombosis with thrombocytopenia syndrome
VITT: vaccine-induced immune thrombotic thrombocytopenia

## ACKNOWLEDGEMENTS

The authors are grateful to the supporting staff of the Forensic Medicine Unit in gathering data. This study received no specific funding.

## CONFLICT OF INTEREST

L.P. is a former small shareholder of AstraZeneca. H.N. is a member of the National Immunization Technical Advisory Group of Finnish Institute for Health and Welfare (THL). H.N. is chair of the WHO Strategic Advisory Group of Experts. THL has until 9/2022 conducted Public-Private Partnerships with vaccine manufacturers and has previously received research funding for studies unrelated to COVID-19 from GlaxoSmithKline Vaccines, Pfizer, and Sanofi Pasteur (including T.N. as an investigator). All other authors declare no financial or non-financial competing interests.

